# Quantifying potential immortal time bias in observational studies in acute severe infection

**DOI:** 10.1101/2025.01.09.25320251

**Authors:** Tom A. Yates, Tom Parks, Peter J. Dodd

**Affiliations:** Institute of Health Informatics, University College London, UK; Division of Infection and Immunity, University College London, UK; Department of Infectious Disease, Imperial College London, UK; Sheffield Centre for Health and Related Research, University of Sheffield, UK

## Abstract

**KEY POINTS:** Immortal time bias exaggerates estimates of treatment efficacy in naïve analyses of observational data. We developed a tool to estimate the extent of this bias. The benefits of giving intravenous immunoglobulin in streptococcal toxic shock syndrome have likely been overstated

**Background:** Immortal time bias is a spurious or exaggerated protective association that commonly arises in naive analyses of observational data. It occurs when people receive the intervention because they survive, rather than survive because they received the intervention. Studies in conditions with substantial early mortality, such as acute severe infections, are particularly vulnerable. The bias can be avoided by fitting the intervention as a time-varying exposure.

**Methods:** We developed IMMORTOOL, an R package accessible via a user-friendly web interface. This tool will estimate the potential for immortal time bias using empiric or assumed data on the distributions of time to intervention and time to event. Assumptions are that no other biases are present and that the intervention does not impact the outcome. The tool was benchmarked using studies presenting both naive analyses and analyses with the intervention fit as a time-varying exposure. We applied IMMORTOOL to a set of influential observational studies that used naive analyses when estimating the impact of polyclonal intravenous immunoglobulin (IVIG) on survival in streptococcal toxic shock syndrome (STSS).

**Results:** IMMORTOOL demonstrated that published estimates suggesting a survival advantage from giving IVIG in STSS are explained, at least in part, by immortal time bias.

**Conclusions:** IMMORTOOL can quantify the potential for immortal time bias in observational analyses. This may help readers interrogate published studies. We do not advocate IMMORTOOL being used to justify naive analyses where the intervention could be fit as a time-varying exposure. To what extent giving IVIG in STSS improves survival remains uncertain.

## Background

Patients may succumb to their disease before a treatment is initiated. In this scenario, observational studies that simply count deaths in the treated and untreated groups will overestimate the benefits of the intervention because patients need to survive long enough to receive treatment. Here, some of any ‘protective’ association will result from people having received the intervention because they survived, rather than from them surviving because they received the intervention.

This phenomenon is immortal time bias (ITB), a problem reportedly first described by William Farr in 1843[1] then rediscovered in the 1970s.[2] Pre-treatment time in the treated group is misclassified because patients are yet to receive treatment (Figure 1). This time period is ‘immortal’ because it is not possible to die prior to treatment and to then receive treatment. Other terms for this type of survival bias include ‘time dependent bias’ and ‘survivor treatment selection bias’.

**Figure 1.**
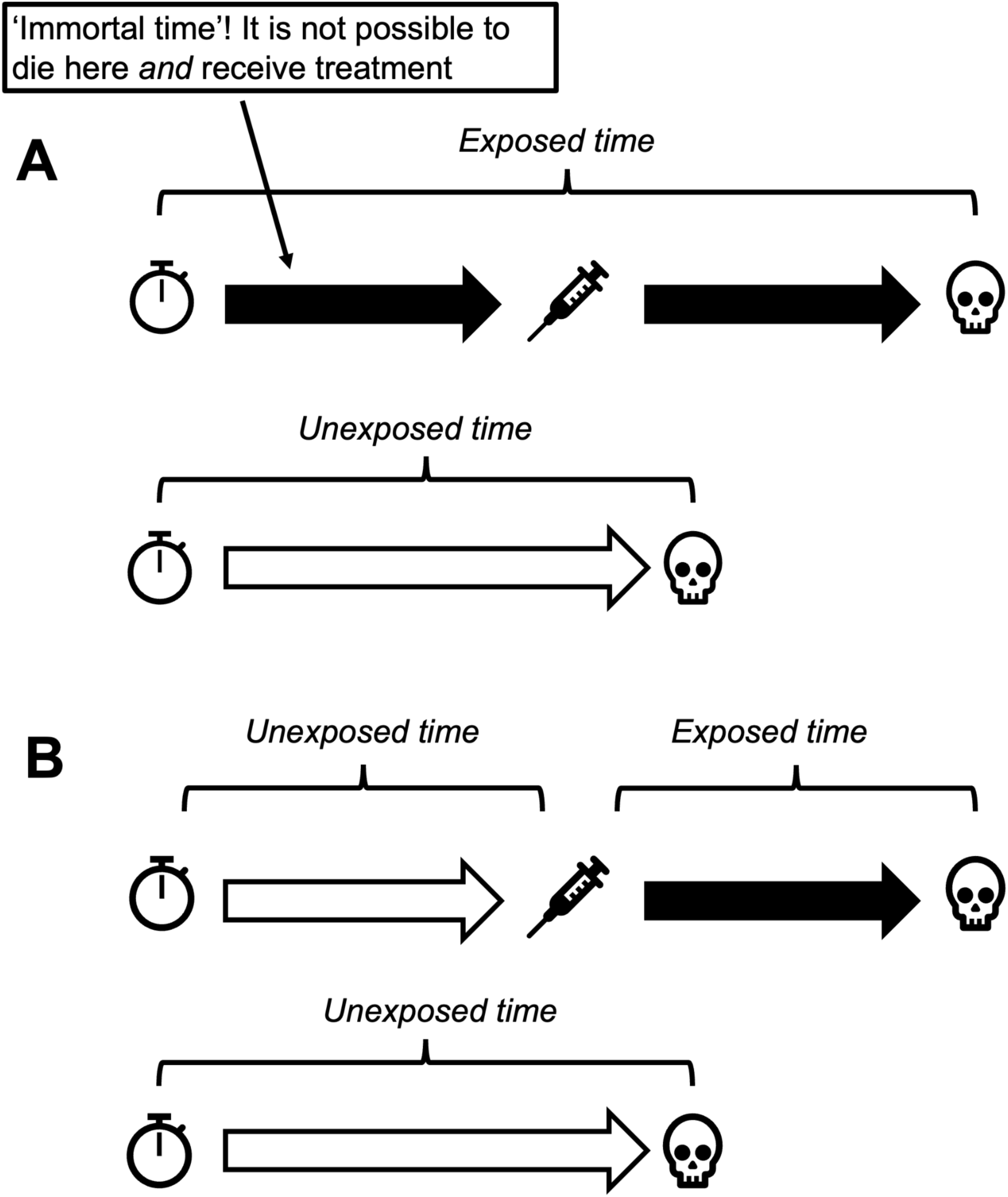
A schematic depicting the attribution of pre-treatment and post-treatment time in the exposed and unexposed groups applying (A) a naive analysis and (B) the correct analysis, with the intervention fit as a time-varying exposure.

Naive analyses of this sort are common despite there being analytical approaches available that result in person time being properly allocated (Table 1).[3] The two best approaches to avoiding misassignment of person time are analysing the intervention as a time-varying exposure and designing the study so that follow up time in treated and untreated individuals is aligned.

**Table 1.** Some analytical approaches that are used to address misallocation of person time.

| Solution | Advantages | Disadvantages |
| --- | --- | --- |
| Optimal solutions |  |  |
| Fit intervention as a time-varying exposure | Addresses ITB | Requires granular data on both time to intervention and time to event<br>Requires authors to have some statistical modelling skills |
| Design study or analysis so that follow up time in treated and untreated group are aligned[20,24] | Addresses ITB and related biases | Not always possible or straight forward |
| Occasionally reasonable solutions |  |  |
| 'Landmark analysis'<br>(see text) | Computationally easy<br>Can often be applied in datasets lacking granular data on time to intervention or time to event | Throws away data<br>Produces estimates conditional on survival to landmark time, which may not be generalisable to individuals treated early |
|  | Addresses ITB if appropriate<br>'landmark time' chosen |  |
| Problematic fixes |  |  |
| Exclude early events | Computationally easy<br>Can often be applied in datasets lacking granular data on time to intervention or time to event<br>May attenuate ITB | Some residual ITB is likely<br>Throws away data<br>Produces estimates conditional on survival to the timepoint chosen, which may not be generalisable to individuals treated early |
| Exclude untreated individuals who die early | Computationally easy<br>Can often be applied in datasets lacking granular data on time to intervention or time to event<br>May attenuate ITB | Throws away data<br>Not comparing like with like, as treated patients who die early will be counted among the deaths - results may be hard to interpret |

Another approach - ‘landmark analysis’ - is widely used.[4] Here, follow up in both the treated and the untreated group starts at a time by which most treatment has been administered (landmark time), with treatment status fixed at that point. This means individuals that are subsequently treated will be counted among the untreated group. If an appropriate landmark time is chosen, ITB is largely addressed. However, the approach is inefficient, disregarding information on events occurring prior to landmark time.[4] Furthermore, it generates estimates conditional on survival to landmark time. These estimates may not be generalisable to the overall population being considered for treatment, because the treatment decision is mostly taken at an earlier point in the disease process. For example, antibiotics may be less effective if administered late.

Various other, mostly problematic, ‘fixes’ to address ITB are also commonly applied (Table 1). Rather than being a consequence of ITB itself, the shortcomings of these approaches principally reflect different treatment of exposed and unexposed groups. For example, time zero may not be aligned, or early deaths may be discounted in only one of the two groups. Such decisions mean that investigators are not comparing like with like, e.g. comparing people at different stages of their illness or comparing survivors with an unselected group.

The extent to which ITB impacts estimates of treatment effect will vary. If patients typically receive the intervention at a time-point that is earlier than most observed deaths, then the bias will be small. Whilst our focus here is mortality, ITB can also occur where other endpoints preclude receiving the intervention. For example, medications that cannot be given in acute kidney injury may appear to protect against renal impairment because they will be prescribed more often to patients whose renal function is preserved.

Here, we describe the design and validation of a simple tool that can be used to assess the extent to which ITB might bias the results of observational studies that do not use methods to account for immortal time, or which use commonly applied but problematic methods, such as landmark analysis. Whilst the tool can be applied to any observational study, the studies that motivated this work were observational analyses of interventions in acute severe infections where early mortality is high. Such analyses are particularly vulnerable to ITB, with naive analyses often generating results that suggest interventions have implausibly large benefits.

Our tool only addresses bias resulting from misclassification of person time. A set of related forms of selection bias that can occur despite the intervention being fit as a time-varying exposure are mentioned in our conclusions section.

## Methods

### Description of the tool

We developed an R package called IMMORTOOL, including a web-browser interface, which will assess the potential for ITB to affect study results. The tool can fit Weibull mortality and exposure hazards using the data on cumulative outcomes by time that are typically reported in studies. The tool accounts for competition between mortality and exposure. Plots allow assessment of the plausibility of fits obtained. Under the assumption that the intervention has no effect, fitted hazards are then used to simulate a cohort with default size 100,000.

Apparent effect sizes are calculated as mortality rate ratios (RRs) using four commonly applied analytical approaches:

a. **Person-time from time zero:** Pretreatment time in the treated misclassified as ‘exposed’ (our default, and the most commonly applied naive approach);
b. **Exclude early events and do not reset clock:** As above, but discounting deaths in both the treated and untreated groups that occur before a prespecified follow up time, leaving time zero unchanged;
c. **Exclude early events and reset clock:** As above, but discounting deaths in both the treated and untreated groups that occur before a prespecified follow up time, and also resetting time zero to be that same time point;
d. **Landmark analysis:** As above, but defining exposure status at the landmark time, i.e. individuals treated after landmark time are retained in the untreated group.

These approaches are defined formally in the Appendix and some advantages and disadvantages given in Table 1. For each approach, the RR is calculated as the ratio between the total deaths over total person-time for those exposed vs those unexposed. Finally, approximate confidence intervals are calculated using the observed number of deaths and the expected fraction of deaths occurring in the exposed vs unexposed group (see Appendix).

### Benchmarking

To assess the performance of our tool, we used a set of infection-related observational studies that reported the same treatment effects using methods that accounted for ITB as well as methods that did not account for ITB. Detailed descriptions of each of these studies can be found in the Appendix.

Two of these studies undertook both naive analyses, but also fitted the intervention as a time-varying exposure, thereby addressing ITB. The first described mortality in people with *Staphylococcus aureus* bacteraemia who had survived for 48 hours after the date of the first positive blood culture.[5] The analysis compared mortality in patients who received or did not receive a Computed Tomography - Positron Emission Tomography (CT-PET) scan to screen for deep foci of infection. The second study described mortality in people admitted to intensive care beds with severe H1N1 influenza, comparing mortality in people who received or did not receive oseltamivir, an antiviral drug.[6,7] The authors of this influenza study also provided a landmark analysis.

In both these studies, correct allocation of person time substantially attenuated the protective association reported. It is important to note that observational analyses properly accounting for ITB may not estimate the true effects of the intervention, as confounding by indication and other biases may result in either an underestimate or an overestimate of the true association.[8] Randomized controlled trials (RCTs) of both interventions are ongoing (NCT04381936, NCT02735707, NCT05137119).

The third study described the rate with which people admitted to hospital with COVID-19 progressed to mechanical ventilation or death.[9] The analysis compared the incidence of this composite endpoint in people who received or did not receive the drug hydroxychloroquine. The authors presented both a naive analysis and a landmark analysis, but did not include an analysis with the intervention analysed as a time-varying exposure. Both this observational analysis and subsequent RCTs suggested that the intervention - hydroxychloroquine as a treatment for COVID-19 - offered either no benefit or was harmful.[10]

### Application to a clinical problem: IVIG in STSS

We next used our tool to assess the likely impact of ITB on observational studies in acute infection that did not model the intervention as a time-varying exposure. Here, our focus was on observational studies of polyspecific intravenous immunoglobulin (IVIG) in patients with streptococcal toxic shock syndrome (STSS).

We selected this example because STSS carries very high early mortality. IVIG is an expensive pooled blood product and, in many settings, not immediately available. As such, the risk of meaningful ITB seemed high. The only RCT of IVIG in STSS enrolled 18 patients, so cannot exclude either substantial benefit or substantial harm.[11]

We took our data from a systematic review of observational studies describing mortality in patients with STSS who either received or did not receive IVIG.[12] The review reported 30 day mortality in patients, all of whom had received adjunctive clindamycin. Data in this subgroup was not reported in all of the primary papers.

Three of these studies applied a naive analysis.[13–15] One study excluded untreated (but not treated) individuals who died within the first twelve hours.[16] We applied IMMORTOOL to the first three studies but not to the study excluding early deaths in the untreated. Whilst excluding early deaths in the untreated might be expected to attenuate ITB, it comes at the cost of creating groups that are not directly comparable – i.e. all treated individuals compared to untreated individuals who had survived for at least twelve hours. The net impact of this strategy is not something that IMMORTOOL is able to predict.

Only one of the remaining studies reported time to receipt of IVIG, stating that this was mostly initiated ‘during the first day of onset of illness’.[15] In our primary analysis, for all studies, we assumed that, in the treated group, 50% received the intervention within 12 hours. With only one time point - 30 day mortality - provided in the clindamycin treated STSS population, we used external data to parameterise the distribution of time to death.[17] In our primary analysis, we assumed that all deaths occurred within 8 days and that half of all deaths occurred within the first 48 hours. To emulate the original review, we pooled results using random effects meta-analysis.

In sensitivity analysis, we varied these assumptions to generate a ‘low ITB scenario’ (50% of all IVIG given within 6 hours, and 33% of all deaths occurring within the first 48 hours) and a ‘high ITB scenario (50% of all IVIG given within 24 hours, and 66% of all deaths occurring within the first 48 hours).

### Code and tool availability

We have made IMMORTOOL available as an R package, with a ShinyApp interface, so readers can undertake their own analyses. The underlying code, including code to reproduce all results reported in this paper, are available at https://github.com/petedodd/IMMORTOOL. Online access to the Shiny interface is available at https://petedodd.github.io/IMMORTOOL-live/. Note, in some browsers, the interface will take a few minutes to load.

### Ethical approvals

All analyses presented here used data in the public domain. Ethical approval to undertake these analyses was, therefore, not required.

## Results

### Benchmarking

Results from the three papers we used for benchmarking are presented in Table 2. Where possible, we present the results from the naive analysis as well as results treating the intervention as a time-varying exposure. Where papers presented both crude and adjusted estimates, we present their primary adjusted estimates. These are presented alongside the estimates provided by IMMORTOOL. The IMMORTOOL results are an estimate of the intervention effect that might be expected purely as a result of ITB, under the assumption that the intervention had no effect and that no other biases were at play.

**Table 2.**
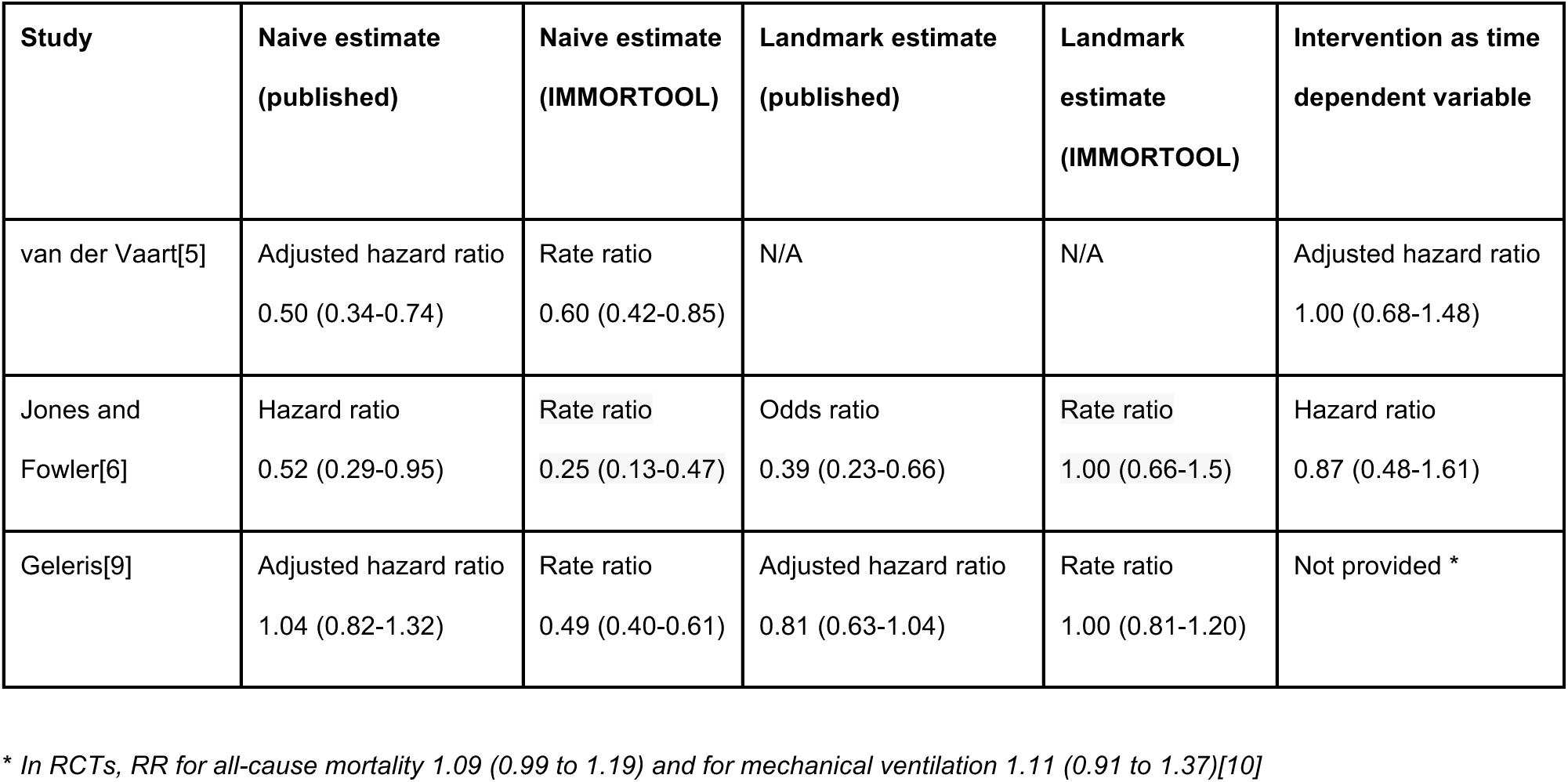
Published results from the three ‘benchmarking’ studies alongside estimates from IMMORTOOL.

| Study | Naive estimate<br>(published) | Naive estimate<br>(IMMORTOOL) | Landmark estimate<br>(published) | Landmark<br>estimate<br>(IMMORTOOL) | Intervention as time<br>dependent variable |
| --- | --- | --- | --- | --- | --- |
| van der Vaart[5] | Adjusted hazard ratio<br>0.50 (0.34-0.74) | Rate ratio<br>0.60 (0.42-0.85) | N/A | N/A | Adjusted hazard ratio<br>1.00 (0.68-1.48) |
| Jones and<br>Fowler[6] | Hazard ratio<br>0.52 (0.29-0.95) | Rate ratio<br>0.25 (0.13-0.47) | Odds ratio<br>0.39 (0.23-0.66) | Rate ratio<br>1.00 (0.66-1.5) | Hazard ratio<br>0.87 (0.48-1.61) |
| Geleris[9] | Adjusted hazard ratio<br>1.04 (0.82-1.32) | Rate ratio<br>0.49 (0.40-0.61) | Adjusted hazard ratio<br>0.81 (0.63-1.04) | Rate ratio<br>1.00 (0.81-1.20) | Not provided * |

\* In RCTs, RR for all-cause mortality 1.09 (0.99 to 1.19) and for mechanical ventilation 1.11 (0.91 to 1.37)[10]

In the same table, we present the results of landmark analyses by Jones and Fowler[6] and by Geleris *et al*[9] alongside estimates, applying the same landmark time, provided by IMMORTOOL. The IMMORTOOL results, again, represent an estimate of the intervention effect that might be expected purely as a result of residual ITB, under the assumption that the intervention had no effect and that no other biases were at play.

### IVIG in STSS

Estimates of the effects of IVIG on mortality in STSS are presented in Figure 2. All three observational studies applied a naive analysis, and the data presented here are unadjusted for potential confounders. Also presented in Figure 2 are estimates from IMMORTOOL, under various assumptions about the distributions of time to event and time to intervention, plus the estimate from the single underpowered RCT.

**Figure 2.**
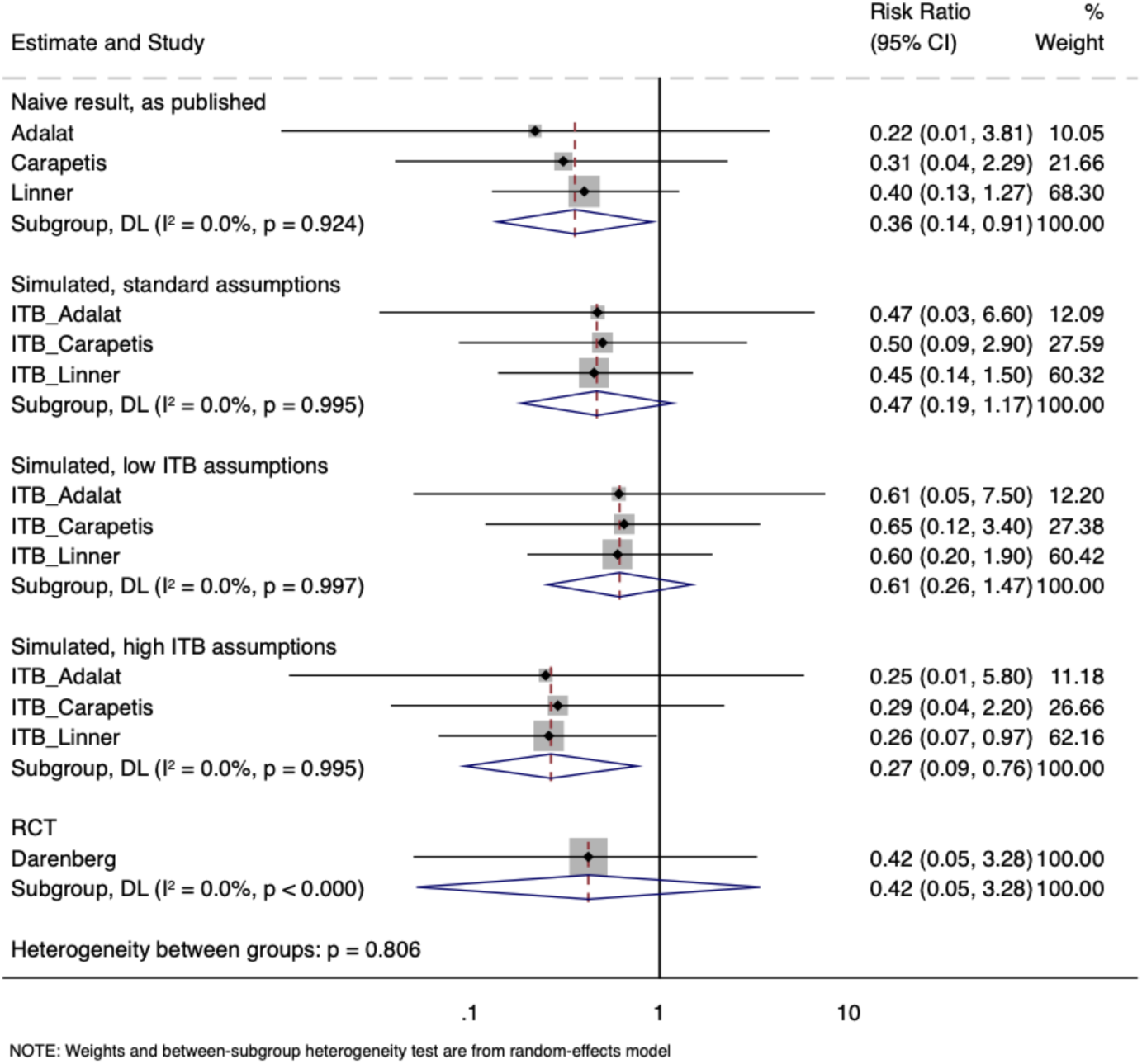
A forest plot containing the results reported by three observational studies estimating a risk ratio for 30 day mortality in patients with STSS according to whether they received IVIG; the results of three sets of IMMORTOOL simulations, estimating the results expected as a result of ITB, if the intervention has no effect on the outcome; and the results of the small RCT. The standard simulation assumptions had 50% of those receiving the intervention receiving it within 12 hours, and half of all deaths occurring within 48 hours. The low ITB assumptions had 50% of those receiving the intervention receiving it within 6 hours, and 33% of all deaths occurring within 48 hours. The high ITB assumptions had 50% of those receiving the intervention receiving it within 24 hours, and 66% of all deaths occurring within 48 hours.

## Conclusions

Inappropriate analyses of observational studies, that fail to account for immortal time, can result in the protective effects of interventions being overestimated. This is a particular risk in conditions with substantial early mortality, such as acute severe infections. By fitting to reported data and simulating a study with a true null effect, IMMORTOOL can calculate the likely extent of this bias. In doing so, it makes two key assumptions. The first, that the intervention has no effect on the outcome, is often true. The second, that no other biases are at play, is rarely true.

In both the CT-PET and influenza studies, IMMORTOOL identified the potential for substantial ITB in the naive analyses, which was apparent when comparing these to the results fitting the intervention as a time-varying exposure. IMMORTOOL also suggested the potential for significant ITB in the hydroxychloroquine study. That the naive estimate in that study suggested no benefit – in keeping with data from randomized controlled trials – may suggest that a second bias, most likely residual confounding by indication, offset the ITB. Here, we did not have results fitting the intervention as a time-varying exposure to directly compare to results from the naive analysis.

Clearly, other biases may attenuate or exaggerate naive effect estimates. In smaller studies, the play of chance will also result in naive estimates that do not align with those predicted by IMMORTOOL. Our benchmarking exercise does, however, suggest that the tool will highlight situations in which substantial ITB is likely.

The landmark analysis performed by the authors of the influenza study is instructive as it resulted in an estimate of the treatment effect that was further from the estimate with the intervention fit as a time-varying exposure than the estimate from the naive analysis. This is likely because the approach throws away data, resulting in imprecision.

We used IMMORTOOL to interrogate three influential studies of the association between receiving IVIG and 30 day mortality in STSS.[13–15] Even where conservative assumptions were made regards the distribution of time to intervention and time to event, IMMORTOOL suggested that ITB was leading to exaggerated estimates of the benefits associated with IVIG treatment. Whilst our results suggest that published estimates of the effects IVIG on mortality in STSS overstate the survival advantage because of ITB, we would caution against concluding that the intervention has no benefit. There are reasons that observational studies might underestimate the benefits of this intervention. For example, if IVIG were offered to the sickest STSS patients then a modest benefit might be masked by confounding by indication. Confounding by indication could also operate in the other direction, with IVIG administration associated with prompt STSS diagnosis and proactive management.

The most robust means of answering this important clinical question would be to undertake a large pragmatic multicentre RCT. The acuity of the situation, potential harms associated with administering a blood product, and the significant limitations of the existing evidence base could argue in favour of deferred consent, as has been used in other recent trials in acute severe infection.[18] Deferred consent would likely improve enrolment. However, if the true benefits of IVIG are more modest than had previously been assumed, then this RCT would need to be large.[19] The only previous trial ceased recruitment after randomizing 18 participants because enrolment was too slow.[11]

Our tool can assess likely ITB in specific cases, and includes the impact of sample size on precision. In smaller samples, exaggerated point estimates - in both directions - will be seen more commonly regardless of the extent of ITB. IMMORTOOL can also explore the impact of landmark analysis and of crude approaches to limit ITB, such as not including deaths that occur early.

Importantly, ITB is related to other forms of selection bias, such as prevalent user bias, that are not fully addressed by fitting the intervention as a time-varying exposure (this only addresses misallocation of person time). These biases occur where the intervention is harmful or protective - contrary the assumption made by IMMORTOOL - and where treatment assignment and follow up time are not aligned. Here, the most vulnerable individuals may be selectively depleted from the exposed or unexposed group such that, when follow up begins, the two populations are not comparable. These issues, and methods that can address selection problems beyond misallocation of person time, are discussed elsewhere.[20]

A challenge in applying IMMORTOOL is that study authors often do not provide full data on the timings of the intervention or endpoints. However, readers will often have some idea of the expected distribution of time to death and can test a number of plausible time to intervention distributions.

It is noteworthy that reporting guidelines do not explicitly mandate reporting of the distribution of time to treatment or time to event in observational studies of health interventions. STROBE[21] requires authors to ‘report numbers of outcome events or summary measures over time’, but there is nothing further in the relevant STROBE extensions RECORD[22] or RECORD-PE.[23] Updating these reporting guidelines to encourage the reporting of these distributions, so readers have the data required to assess risk of ITB, should be considered.

IMMORTOOL enables quantitative assessments of the likely extent of ITB in published observational studies. This may flag studies where the published intervention effect is likely to be over optimistic. Alternatively, it may provide reassurance that ITB is unlikely, meaning published results are not automatically disregarded, although clearly other forms of bias may still explain results.

It is not our intention that the tool be used to justify naive analyses where better analyses are possible. However, sometimes data do not allow for the intervention to be fit as a time-varying exposure. For example, sufficiently granular data on time to intervention or time to event may not be available. Here, investigators may find IMMORTOOL helpful in deciding whether to proceed with a crude or naive analysis. Where possible, important questions regards the relative efficacy and safety of interventions should be answered in randomised controlled trials.

## Supporting information

Technical appendix

## Data Availability

https://petedodd.github.io/IMMORTOOL-live

## Acknowledgements

We are grateful to Dr Mark Jones and Prof Rob Fowler for answering our queries about their paper exploring the impact of immortal time bias on estimates of the efficacy of oseltamivir in severe influenza.

## Patient consent

As this was a secondary analysis of published data, patient consent was not required.

