## Supplementary material for "Quantifying potential immortal time bias in observational studies in acute severe infection": Technical appendix

### Fitting to data

The package includes utilities to fit Weibull hazards of mortality and exposure (i.e. receiving treatment) in a competing hazards framework. The input data is a set of times and corresponding fractions of the initial cohort that have cumulatively experienced the event by that time.

Let $m_i$ with $i = 1, \cdots, N_m$ be the times at which corresponding fractions $M_i$ of the initial cohort have died. Because we assume that death is independent of treatment, for a Weibull hazard we have

$$M_i = \exp\left(-\ (m_i/L_m)^{k_m}\right)$$

which rearranges as

$$\log(-\ \log(M_i)) = k_m \log m_i - k_m \log L_m.$$

The Weibull shape $k_m$ and scale $L_m$ parameters for mortality can be obtained from a linear regression with this transformed data: the slope yields the shape parameter directly, and the scale parameter can then be obtained from the intercept.

Because exposure can only happen to individuals who have not died, the probability density function, $p(t)$, for exposure happening in a small interval around time $t$ is

$$p(t) = \frac{k_e}{L_e}\left(\frac{t}{L_e}\right)^{k_e - 1} \exp\left(-\ (t/L_e)^{k_e}\right) \times \exp\left(-\ (t/L_m)^{k_m}\right), \qquad [*]$$

which is the probability of avoiding deaths or exposure to time $t$ and then experiencing exposure, when the exposure hazard is Weibull with shape parameter $k_e$ and scale parameter $L_e$.

The cumulative probability of someone in the initial cohort being exposed by time $t$ is therefore

$$P(t) = \int_0^t d\tau\, p(\tau).$$

To fit to data $x_i$ with $i = 1, \cdots, N_e$ denoting the times at which corresponding fractions $X_i$ of the initial cohort have been exposed, we numerically minimise the sum of squared error (SSE)

$$SSE = \sum_{i=1}^{N_e} \left(X_i - P(x_i)\right)^2$$

over exposure hazard parameters $k_e$ and $L_e$ once mortality parameters have been fixed by data.

There is also an option to fit to data where the fractions $X_i$ are interpreted as fractions of the cohort ever exposed who have been exposed by time $x_i$. In this case, $\bar{P}(t) = P(t)/P(\infty)$ (the cumulative probability of exposure conditional on exposure) is used instead of $P(t)$ in the above calculations.

#### Approximation for numerical stability

In calculating the SSE, we need the cumulative distribution given by the integral of equation [*], which we calculate numerically. However, when $k_e < 1$ this can cause problems for the numerical integration close to 0, and so for $k_e < 1$ and $T/L_e < 1/200$ we use the series expansion of this integral

$$P(T) = \int_0^T dt\, p(t) = \sum_{l=0}^{\infty} \frac{(-1)^l}{l!} \left(\frac{L_e}{L_m}\right)^{k_m} \gamma\left(1 + \frac{lk_m}{k_e}, \left(\frac{T}{L_e}\right)^{k_e}\right)$$

up to $l = 1$ (where $\gamma(a, x)$ represents the lower incomplete gamma function).

### Simulation

Once time-to-event distributions have been obtained for death, exposure, and loss to follow-up (LTFU), a synthetic cohort of default size 100,000 is created with times of death ($t_m$), exposure ($t_e$), and LTFU ($t_l$) independently sampled from corresponding distributions. Time-to-event distributions can be Weibull distributed, with parameters derived via the above fitting procedure, but could also be any other type of distribution for which a pseudo-random number generator exists in R, with parameters derived from separate analysis or simply posited by the user. LTFU is not explored here and simulations assume an exponential distribution with 100 year timescale..

### Effect size calculations

Using a synthetic cohort simulated as described above, total event counts and person-time are used to compute mortality rates. Rate ratios (RRs) are calculated as ratios of mortality rates in those classed as exposed over those not exposed. By default, within the specified simulation time horizon $T$, individuals are coded as having died if $t_m < T$ and $t_m < t_l$; exposed if $t_e < T$ and $t_e < t_l$ and $t_e < t_m$ Four variant approaches are are used for these calculations:

a) **Person-time from time zero.** Person-time $PT = min(t_m, t_l, T)$, i.e. time measured from 0. Exposure status coded as in default, i.e. exposure status at final time definitive.
b) **Exclude early events and do not reset clock.** A special time $T_{exc}$ is introduced. Those dead ($t_m < T_{exc}$) or LTFU ($t_l < T_{exc}$) before this time are dropped from the synthetic cohort, but exposure remains coded as in default, i.e. final exposure status is definitive. Person-time is measured from 0, i.e. $PT = min(t_m, t_l, T)$.
c) **Exclude early events and reset clock.** A special time $T_{exc}$ is introduced. Those dead ( $t_m < T_{exc}$) or LTFU ($t_l < T_{exc}$) before this time are dropped from the synthetic cohort, but exposure remains coded as in default, i.e. final exposure status is definitive. Person-time is measured from $T_{exc}$ , i.e. $PT = min(t_m, t_l, T) - T_{exc}$ .

d) **Landmark analysis.** A special time $T_{landmark}$ is introduced. Those dead ($t_m < T_{landmark}$) or LTFU ($t_l < T_{landmark}$) before this time are dropped from the synthetic cohort. Those exposed after $T_{landmark}$ are recoded as not exposed (i.e. only those with $t_e < min(T_{landmark}, t_m, t_l, T)$ are coded as exposed: exposure status at $T_{landmark}$ is definitive). Person-time is measured from $T_{landmark}$, i.e. $PT = min(t_m, t_l, T) - T_{landmark}$.

As described above, the effect size under these variants is then calculated as

$$RR = \frac{\sum_{j \in E} 1_j(died) / \sum_{j \in E} PT_j}{\sum_{j \in \bar{E}} 1_j(died) / \sum_{j \in \bar{E}} PT_j}$$

where $E$ is the set of synthetic cohort individuals coded as exposed, $\bar{E}$ is the set of cohort individuals coded as not exposed, and $1_j(died)$ is an indicator function for whether individual $j$ is coded as having died.

### Confidence interval calculations

We calculate confidence intervals for specified study size using standard approximations.
Let $a$ be the number of deaths in the exposed and $b$ the number of deaths in the unexposed, and define a factor $F = \exp\left(1.96 \times \sqrt{\frac{1}{a} + \frac{1}{b}}\right)$.
The 95% confidence interval upper and lower bounds for an incidence rate ratio are given approximately as

$$RR_{upper} = RR \times F,$$
$$RR_{lower} = RR / F.$$

In practice, we are using the observed number of deaths $N$ in a study, and taking $a = fN$ and $b = (1 - f)N$, where $f$ is the fraction of deaths expected to occur in those exposed calculated from the simulation.

### Studies used for benchmarking

#### van der Vaart

van der Vaart et al.[1] was an observational analysis seeking to estimate the association between receiving a CT PET scan, to look for a deep focus of infection, and 90 day all-cause mortality in 476 patients with *Staphylococcus aureus* bloodstream infection, who had survived 48 hours following their first positive blood culture. The authors presented both a naive analysis (as a teaching example) and results from a Cox proportional hazards model in which receipt of CT PET was fit as a time-varying exposure.

Time zero in this study was the date of the first positive blood culture. Over follow up, 178 study participants (37.4%) received a CT PET scan, at a median of 9 days (interquartile range 6 - 13 days). There were 25 (5.3%), 102 (21.4%) and 147 (30.9%) deaths by day 7, 30 and 90, with all deaths in the first week occurring prior to CT PET scan. See Figure S1 for fit.

The analytical approach we modelled here was ‘Exclude early events and do not reset clock’, with a *Tearly* of 2 days (48 hours) applied. This is output ‘b’ from IMMORTOOL (see R package vignette).

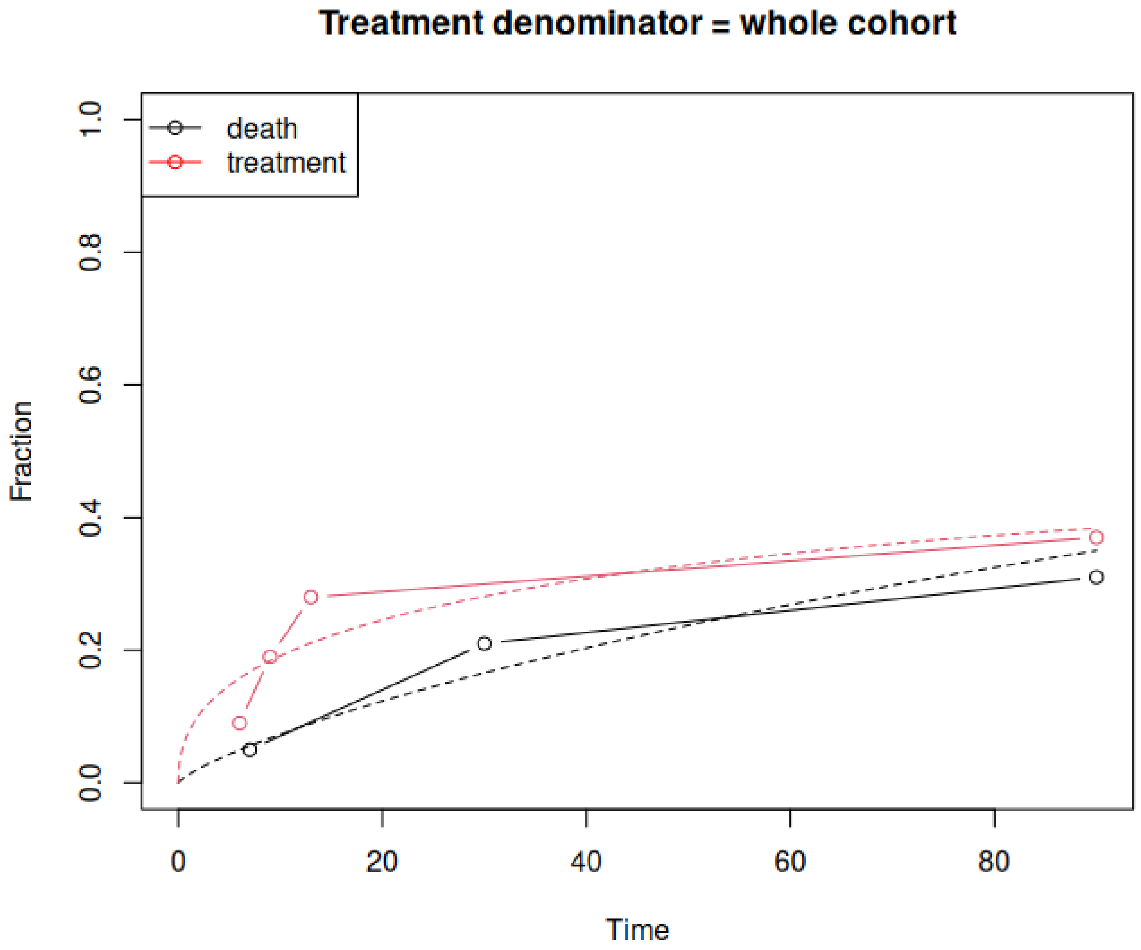

***Figure S1.*** *IMMORTOOL fit for van der Vaart et al.*

#### Jones and Fowler

Jones and Fowler[2] was an observational analysis exploring the association between receipt of oseltamivir and mortality in 578 critically ill patients hospitalised with pandemic H1N1 influenza. As a teaching example, the authors present a naive analysis, results from a Cox proportional hazards model with the intervention fit as a time-varying exposure, and a landmark analysis.

Time zero was admission to the intensive care unit. Over follow up, 540 (93.4%) received oseltamivir and 117 (20.2%) died. Median time to treatment was 0.62 days (range 0 - 45 days). Seven deaths occurred within 24 hours of admission to the intensive care unit with most deaths occurring within 12 days. Granular data on time to intervention and time to event over days 1-12 are provided in an appendix to the Health Technology Assessment and, with the timings of late deaths not specified, we assumed that all deaths had occurred by day 90.[3] See Figure S2 for fit.

The analytical approaches we modelled here were a naive analysis and a landmark analysis applying a *Tlandmark* of 1 day (24 hours). These are outputs 'a' and 'd' from IMMORTOOL (see R package vignette).

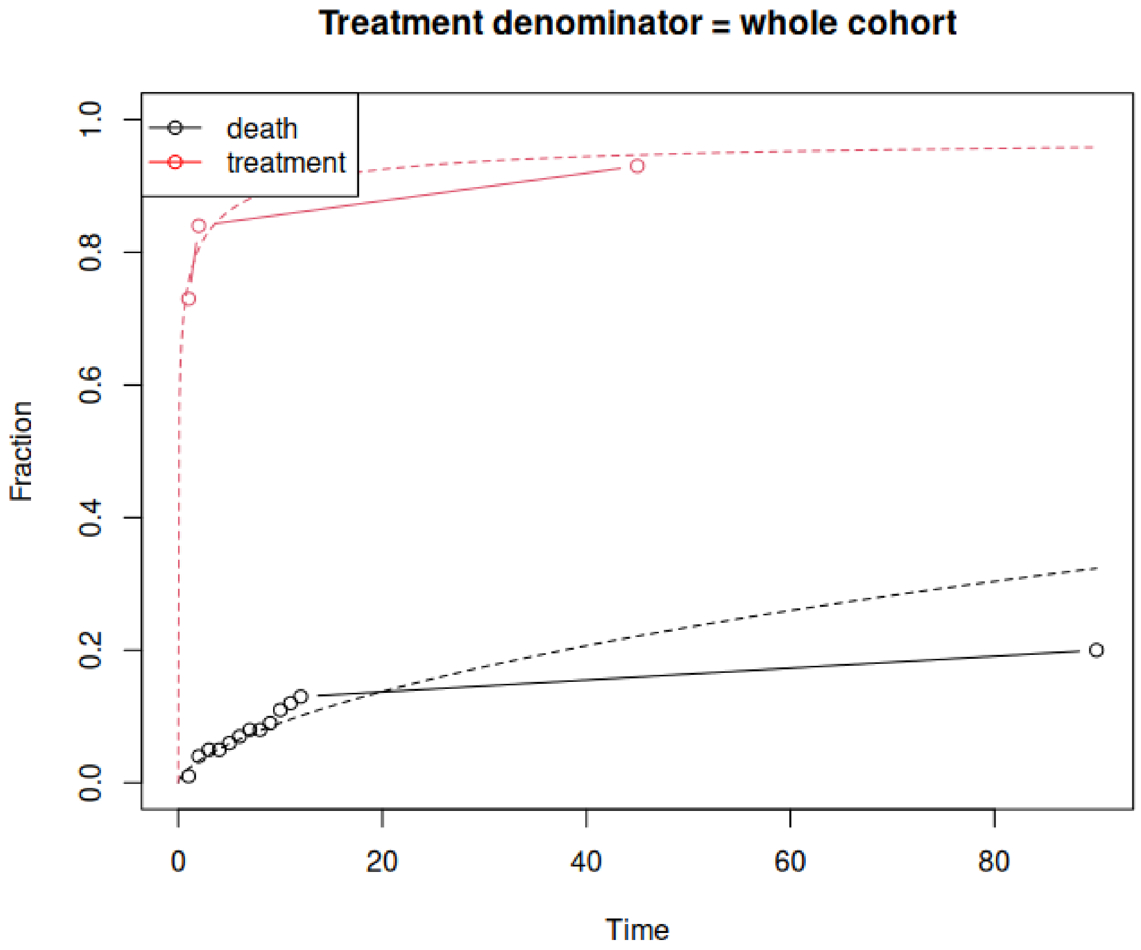

***Figure S2.*** *IMMORTOOL fit for Jones and Fowler*

#### Geleris

Geleris et al.[4] was an observational analysis exploring the association between treatment with hydroxychloroquine and a composite primary endpoint of progression to mechanical ventilation or death in 1376 people hospitalised with COVID-19. The authors presented a naive analysis and also various landmark analyses.

Whilst the times presented in the paper are time since presentation to the Emergency Department, time zero in the paper is set at 24 hours (1 day) after arrival. Over follow up, 881 participants (58.9%) received hydroxychloroquine. Among those treated, 45.8% received the drug in the 24 hours between their presentation to the emergency department and the start of study follow-up, and 85.9% received it within 48 hours after presentation to the emergency department. To input this distribution into IMMORTOOL, the 24 hour time point (time zero) was inputted as 0.1 days and the 48 hour time point as 1 day. Data presented in the paper's supplementary appendix suggests that all treatment was started within 10 days of presentation to the Emergency Department (9 days after time zero). In total, 346 participants (25.1%) met the primary endpoint, with the Kaplan Meier curve suggesting that this largely happened by 30 days. Looking again at data in the paper's supplementary appendix, assuming that most of the difference in sample size between the naive analysis and the landmark analysis was a result of people meeting the primary endpoint, we estimated that five percent of the overall cohort had progressed to mechanical ventilation or death by 48 hours (2 days) after presentation to the Emergency Department. See Figure S3 for fit.

The analytical approaches modelled here were a naive analysis and a landmark analysis, applying a *Tlandmark* of 1 day (24 hours). These are outputs (a) and (d) from IMMORTOOL (see R package vignette).

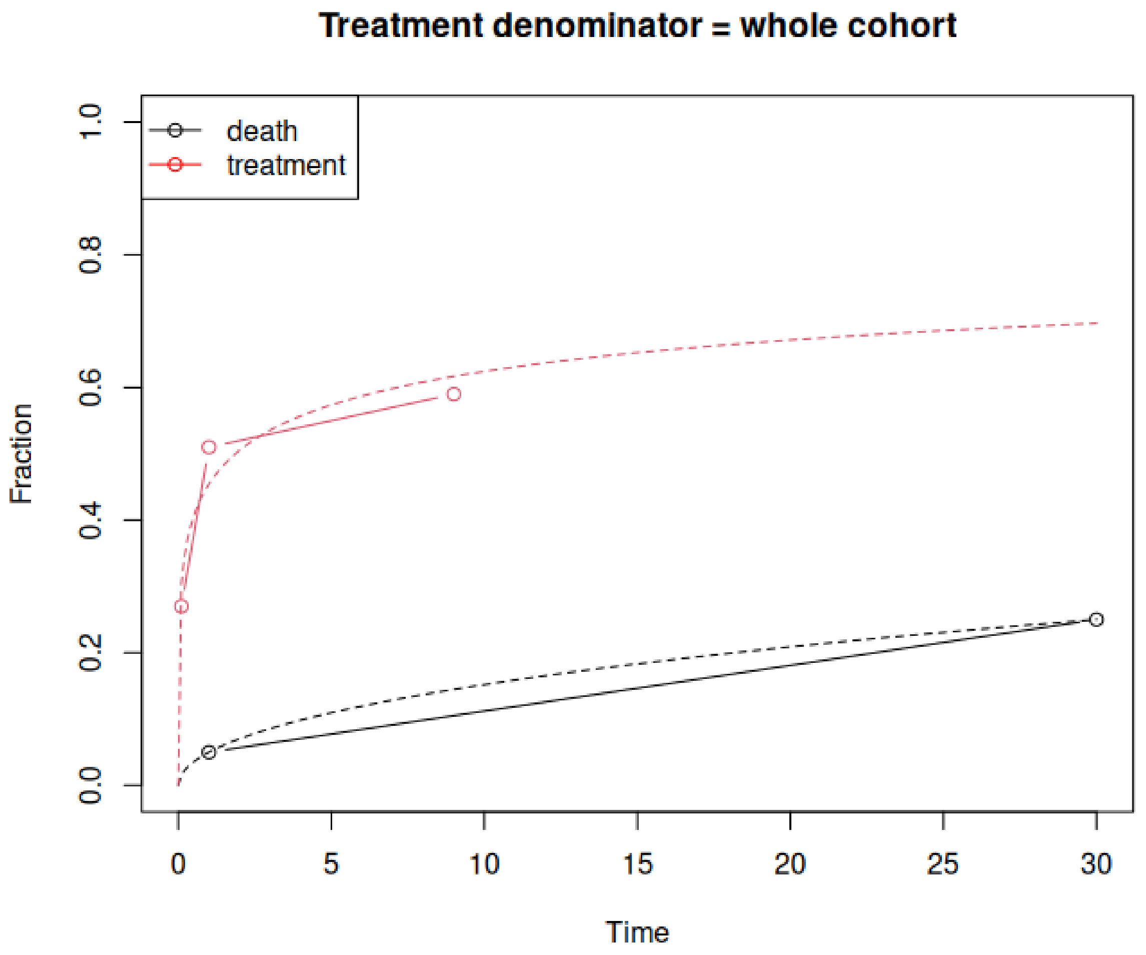

***Figure S3.*** *IMMORTOOL fit for Geleris et al.*
